# Early super-spreader events are a likely determinant of novel SARS-CoV-2 variant predominance

**DOI:** 10.1101/2021.03.23.21254185

**Authors:** Ashish Goyal, Daniel Reeves, Joshua T. Schiffer

## Abstract

The emergence of multiple new SARS-CoV-2 variants, characterized to varying degrees by increased infectivity, higher virulence and evasion of vaccine and infection-induced immunologic memory, has hampered international efforts to contain the virus. While it is generally believed that these variants first develop in single individuals with poor immunologic control of the virus, the factors governing variant predominance in the population remain poorly characterized. Here we present a mathematical framework for variant emergence accounting for the highly variable number of people secondarily infected by individuals with SARS-CoV-2 infection. Our simulations suggest that threatening new variants probably develop within infected people fairly commonly, but that most die out and do not achieve permanence in the population. Variants that predominate are more likely to be associated with higher infectiousness, but also the occurrence of a super-spreader event soon after introduction into the population.

## Introduction

The emergence and subsequent regional predominance of multiple, highly infectious novel SARS-CoV-2 variants in locations across the globe surprised the scientific and public health communities. Several variants are characterized by more than a dozen new mutations often in the genomic region encoding the viral spike protein (1-3).This rate of mutational change surpassed expectations from previous phylogenetic surveys, which identified population sweeps with single new point mutations only after multiple generations of infection (4). Immuno-compromised hosts are a likely source of these variants. Individuals who have impaired cell-mediated or humoral immune function can shed the virus at high viral loads for many weeks, presumably in the relative absence of immune selection pressure (5-8). Yet, it remains unknown why certain variants ultimately predominate.

The spread of new variants has had a dramatic impact on global SARS-CoV-2 epidemiology. The B.1.1.7 variant has higher infectivity and virulence than baseline variants (1, 9), while the B.1.3.5.1 and P.1 variants may have the ability to evade vaccine- and infection-induced immunologic memory (10-12) and may also have higher infectivity and virulence (3, 13, 14). The true abundance of epidemiologically important variants is likely underestimated by sequencing limitations in many global infection hot spots. The B.1.1.7, B.1.3.5.1 and P.1 variants may also undergo further important evolutionary changes over time. Here we perform mathematical modeling to characterize determinants of variant emergence and predominance.

## Results

### Frequent stochastic elimination of new SARS-CoV-2 variants

We previously developed a mathematical model which captures the highly variable secondary transmission pattern of SARS-CoV-2 by fitting to empirically observed distributions of serial intervals within transmission pairs, as well as distributions of number of secondary transmissions from infected people (15-17). The model explains why the majority of infected people do not infect others while a minority are the index case for large super-spreader events (18-20): the exposure contact network of infected people is highly over-dispersed meaning that on rare occasions, an infected person may have dozens of exposure contacts who have the potential to become secondarily infected. Presumably this phenomenon, which is not observed for influenza (21), is due to aerosolization of SARS-CoV-2 within crowded indoor environments in which masking is limited (22). For a super-spreader event to occur, the model projects that the transmitter must also be within the limited time window when they are shedding at a sufficiently high viral load (15). More model details are in the **Methods**.

We adapted this model to assume that new variants emerge from a single infected person. We then ran individual stochastic simulation to assess whether variants are present after 100 days or whether they are extinguished. Simulations were governed by an average effective reproductive number (R_e_) defined as the mean number of secondary infections created per person across the entire population, as well as a value for the “super-spreader parameter” ρ which captures the variability of a gamma-distributed exposure contact network: the value is low (ρ=0.01) for infections such as influenza in which there is low day-to-day and person-to-person variance in the number of exposure contacts, and high (ρ=40) for SARS-CoV-2 (15).

For each parameter set, we ran 1000 simulations. We identified that stochastic burnout of infections is more likely when R_e_ is lower but also when the contact network is highly over-dispersed as with SARS-CoV-2 (**Figure 1a**). This suggests that most highly infectious SARS-CoV-2 variants will die out when generated within a single person, even when R_e_ is quite high.

**Figure 1.**
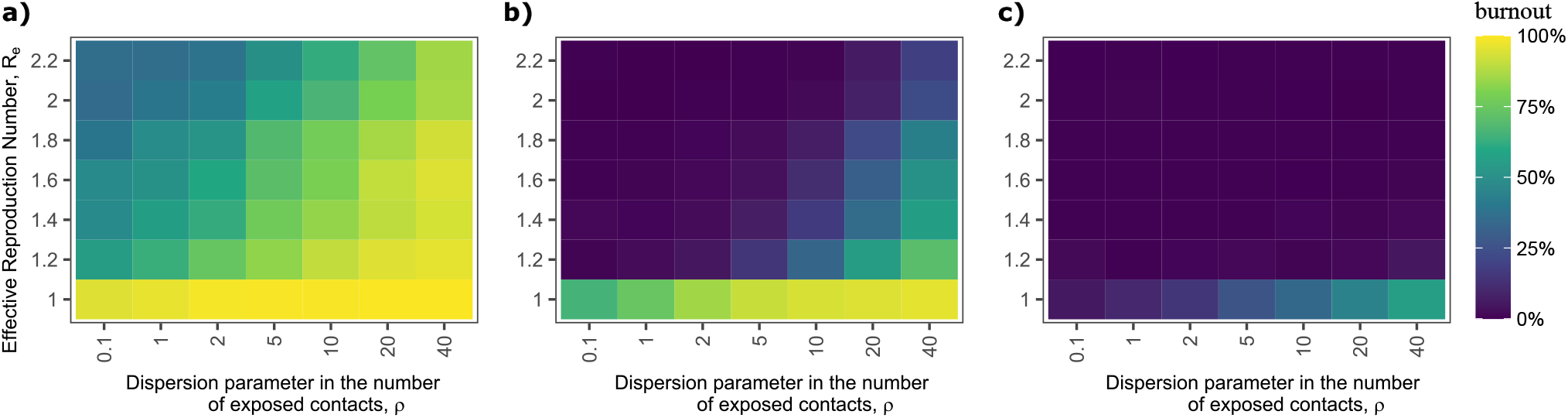
Percentage of new SARS-CoV-2 variant introductions which burn out after introduction into the population. Parameter ρ represents the degree of over-dispersion of exposure contacts. ρ=0.1 is a realistic value for influenza infection and ρ=40 is a realistic value for SARS-CoV-2 infection. The effective reproductive number (R_e_) has varied during the SARS-CoV-2 pandemic depending on the degree of social distancing at a given time. Scenarios assume a) 1, b) 10 and c) 100 initial cases. New SARS-CoV-2 variants have a high percentage of burnout when introduced into the population.

We performed an equivalent analysis with 10 starting infections as might occur if an outbreak of a new variant first spreads in a small household or work cluster. The rate of burnout was still relatively high for low R_e_ and high over-dispersion scenarios but decreased substantially with higher R_e_ values for a given variant (**Figure 1b**). We next performed an analysis with 100 starting infections as might occur with a larger initial super-spreader event. The rate of burnout was low for all R_e_ values and assumptions regarding contact network dispersion (**Figure 1c**). Therefore, although stochastic extinguishment of novel SARS-CoV-2 variants is likely to be common, once roughly 100 cases are established, a variant is likely to continue to expand exponentially in the absence of intensification of non-pharmaceutical interventions (NPIs).

### Timing of SARS-CoV-2 variant emergence is highly variable at realistic effective reproductive numbers

We next evaluated time from first case of a new variant to 1000 cumulative infections in simulations in which stochastic burnout did not occur. We performed 1000 simulations under each assumed value of R_e_. When starting with one infection, we observed a wide variance in time to 1000 infections with an increase in the median time from 23 to 40 days as R_e_ decreased from 2.2 to 1.2 (**Figure 2a**). The variance and median time (19 to 38 days) to 1000 infections were similar when starting from 10 infections (**Figure 2b**) but the median time (7 to 17 days) and variance decreased when starting from 100 infections (**Figure 2c**) again demonstrating that stochastic forces are less important once 100 cumulative infections are reached.

**Figure 2.**
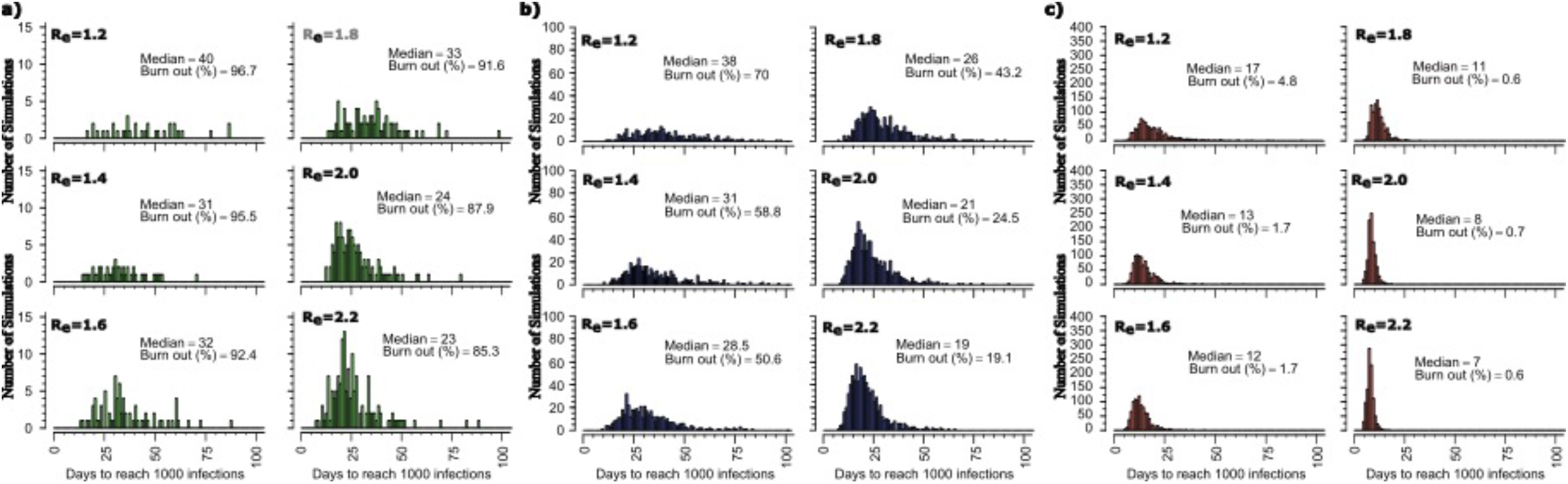
Time to 1000 cumulative infections among SARS-CoV-2 simulations which do not burn out. Scenarios assume a) 1, b) 10 and c) 100 initial cases. Low R_e_ and low number of initial cases is associated with a higher median time to reach 1000 cumulative infections as well as larger variance.

### Early super-spreader events as a predictor of rapid progression to high prevalence of emerging variants

We next examined the timing and number of super-spreader events during these simulations. Super-spreader events were defined as events in which one person infected a minimum of 5 (**Figure 3a**), 10 (**Figure 3b**) or 20 (**Figure 3c**) other people in a day. With each definition and assumed value of R_e_, the timing of the first super-spreader event correlated with time to 1000 cumulative infections. The strength of this correlation increased with the least inclusive definition of a super-spreader event and with higher values for R_e_ (**Figure 3c**).

**Figure 3.**
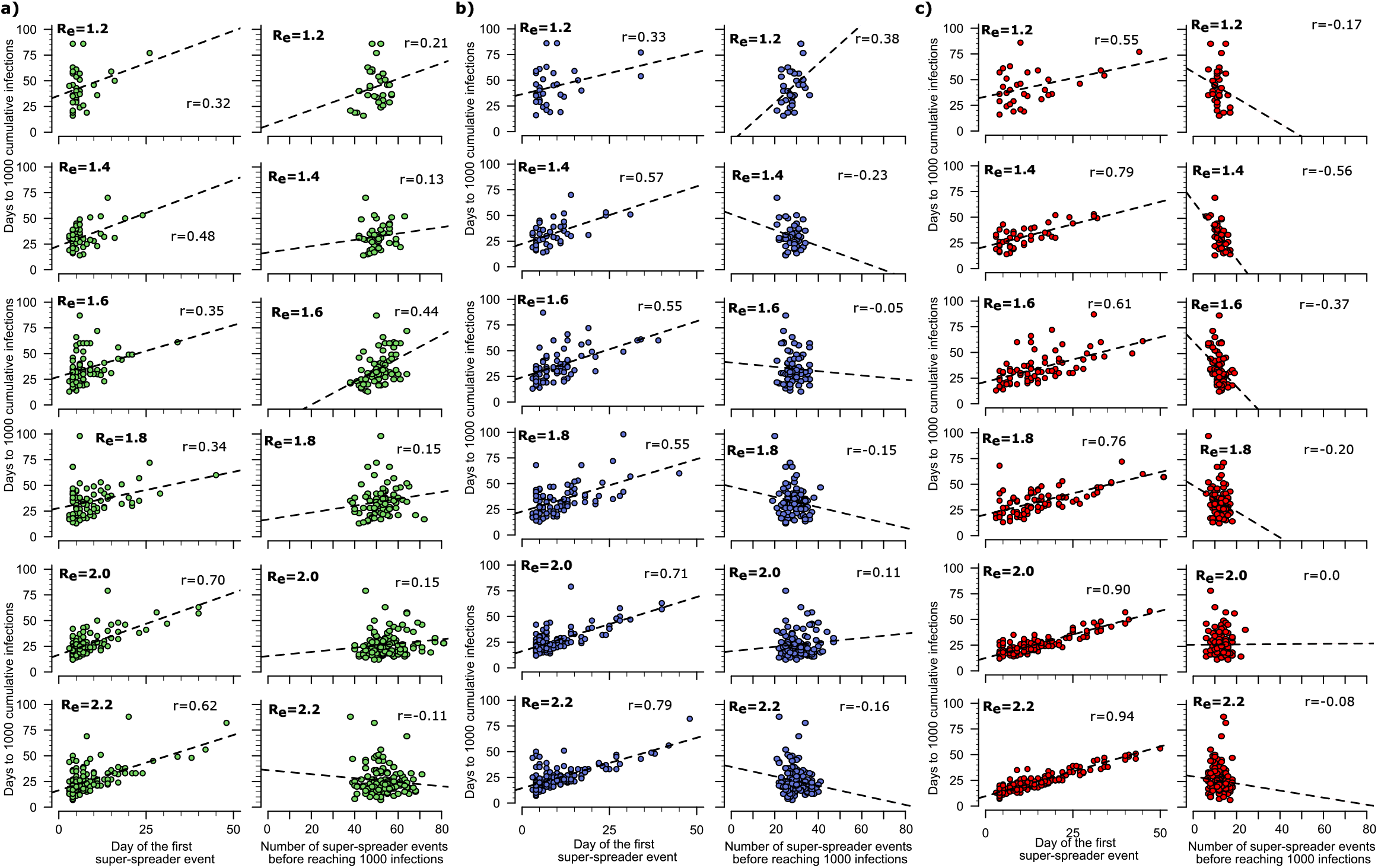
Correlation between timing of super-spreader events and time to 1000 cumulative infections. Scenarios assume definitions of super-spreader events as a) >=5, b) >=10 and c) >=20 secondary infections per day. Early first super-spreader event (left columns) is predictive of more rapid time to 1000 cumulative infections particularly when defined as >=20 infections per day, whereas number of events (right columns) is less predictive.

The number of super-spreader events prior to 1000 cumulative infections correlated positively or not at all with time to 1000 cumulative infections for the more inclusive definitions of super-spreader events (at least 5 or 10 secondary infections, **Figure 3a & b**) across all R_e_ values, signifying that multiple small super-spreader events do not accelerate early epidemic spread. However, the number of super-spreader events correlated negatively with time to 1000 cumulative infections with low to moderate values for R_e_ (1.2-1.8) and less inclusive definitions of super-spreader events (at least 20 secondary infections, **Figure 3c**), meaning that early large super-spreader events are a likely driver of variant predominance under parameter assumptions which are compatible with known features of variant B.1.1.7 (1).

### Increased likelihood of new variant predominance when co-circulating baseline variants have an effective reproductive number less than or equal to one

We next performed analyses assuming a baseline variant at different values for R_e_ (0.8, 1.0 or 1.2) with 1000 baseline infections and assumed that 1% of transmission events result in a new variant. R_e_ for new variants were drawn from a uniform distribution varying from 1.0-2.2 at intervals of 0.2. For each scenario, we performed 100 simulations until 100,000 cumulative infections were generated or until stochastic burnout of all variants occurred. Variants were tracked individually.

Super-spreader events (defined here as >=5 secondarily infected people) were requisite for new variants to reach 1000 cumulative infections in individual simulations. For example, in the case of a baseline variant with R_e_=1.0 and new variant with R_e_=1.2, 95 variants had a super-spreader event, out of which 33 reached 1000 cumulative infections. 905 variants did not have a super-spreader event, and none reached 1000 cumulative infections (p<2.2×10^−16^, Fisher’s exact test). The trend of no variant reaching 1000 cumulative infections in the absence of super-spreader events was observed for all assumed values of new variant R_e_.

When we assumed a baseline variant (starting with 1000 infected at t=0) with R_e_=1.0 (**Figure 4a**), we observed predominance of a new variant in 98 out of 100 simulations (**Figure 4b**). There was a slightly higher likelihood that more infectious variants with higher R_e_ would predominate **(Figure 4b)**. The timing of variant predominance was highly variable (**Figure 4a**). When we performed the same analysis assuming that 0.1% and 0.01% of transmission events result in a new variant, then variant takeover only occurred in 44% and 6% of simulations (**Figure 4c, d**) with more evenly distributed values of R_e_. This suggests that the frequency of variant evolution at the within-host level may determine the characteristics of the predominant strain at the population level.

**Figure 4.**
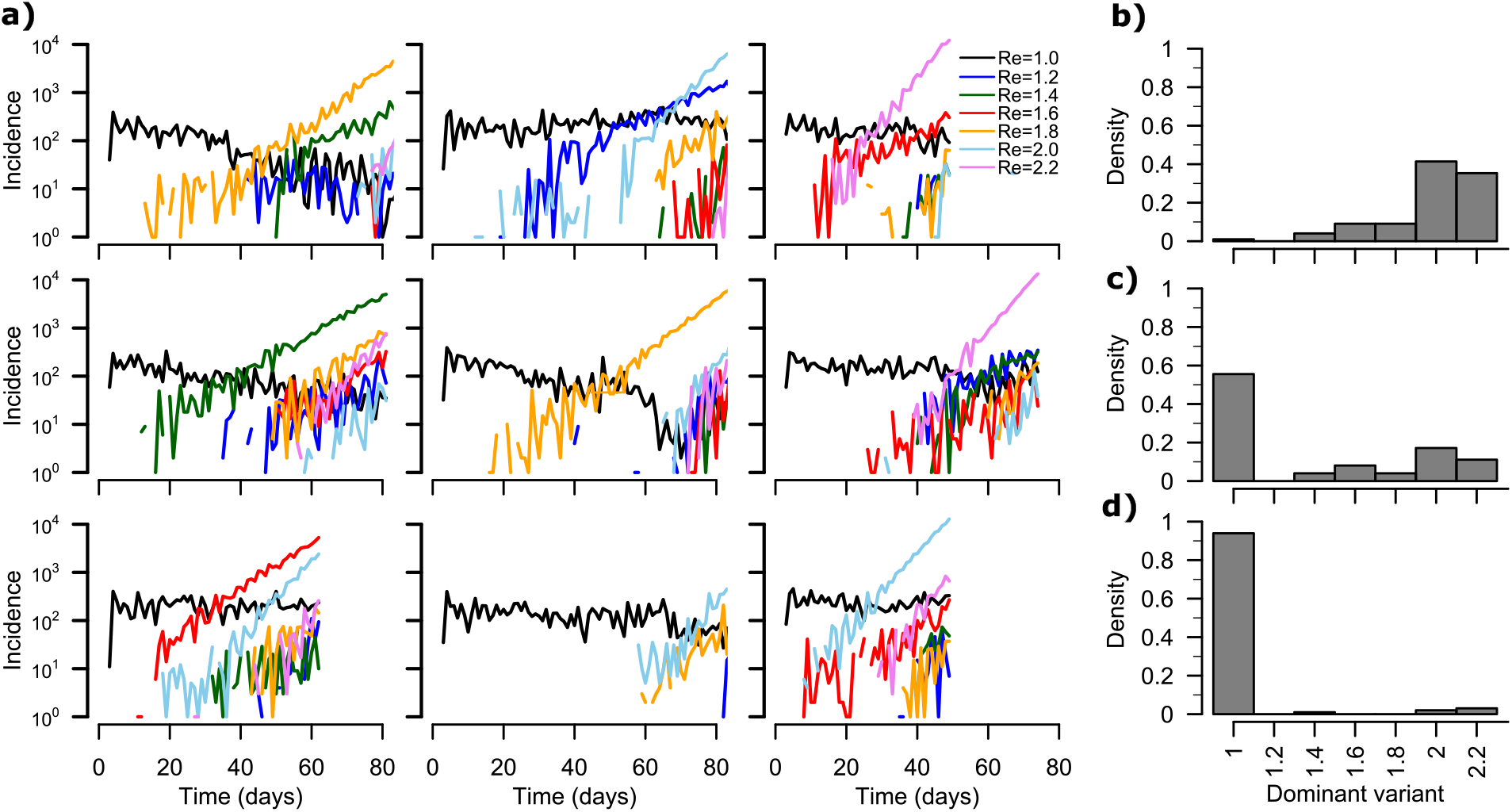
New variant predominance depends on timing of variant introduction, variant effective reproductive number and number of ongoing infections. **a)** Nine example simulations starting with 1000 infections of a baseline variant (black line) with R_e_=1. New variants are colored lines that are randomly generated in 1% of new infections and denoted according to R_e_. The variant which stochastically exceeds 10 cases per day usually predominates though occasionally an infectious variant will expand more rapidly (top middle and left). New variants become much more common as incidence increases. Frequency histograms of R_e_ of new variants with the assumption that **b)** 1%, **c)** 0.1% and **d)** 0.01% of new infections result in new variants with R_e_ of 1.2, 1.4, 1.6, 1.8 or 2.0.

When we performed the analysis using a baseline variant with R_e_=1.2 and 1% of transmission events resulting in a new variant, then variant takeover only occurred in 42 of 100 of simulations, whereas a baseline variant with R_e_=0.8 allowed variant takeover in 100 of 100s simulations with no stochastic burnout of virus. This results explains why new variant predominance is often observed when incidence of the baseline variant is decreasing (1).

### High incidence outbreaks and formation of new variants

In simulations with the baseline variant R_e_=1.0, new high incidence waves of infection were predictably associated with emergence of more new variants some of which ultimately predominated due to higher R_e_ (**Figure 4a**, top and middle left panels). This finding highlights that prevention of new variants is achieved most effectively by avoidance of large waves of infection.

## Discussion

Our simple mathematical model identifies that while higher infectivity is one important predictor of a variant’s ultimate predominance in a population, but there is also a substantial element of bad luck. Stochastic burnout is a common event when a pathogen with a reproductive number between 1 and 2 is introduced into a population (23). The over-dispersed secondary infection rate associated with SARS-CoV-2 further increases the likelihood of stochastic burnout given an equivalent basic reproductive number. This suggests that most new highly infectious variants which emerge from infected individuals never spread substantially in the population. It also raises the more provocative hypothesis that human coronaviruses with pandemic potential such as SARS, MERS and SARS-CoV-2 are introduced into the human population fairly commonly, but that most local outbreaks are avoided due to good fortune alone.

Among variants which do establish a foothold, our model suggests that early large super-spreader events, which are relatively rare at the individual level but become increasingly likely as incidence increases during a local outbreak, may determine which variant is likely to predominate. These events provide a head start for a given variant, bypassing the slower exponential growth phase to a phase of epidemic growth which is more deterministic. Super-spreader events later during an epidemic growth curve are relatively less important for a variant to achieve predominance.

From a public health perspective, our results provide yet another reason to intensely focus non-pharmaceutical interventions (NPIs) on preventing large super-spreader events. This policy prescription includes prohibition of large indoor gatherings, a focus on adequate ventilation in indoor work environments and schools, and enforcement of highest quality masks (K95 or N95) in circumstances where group exposures cannot be avoided (17). Prevention of super-spreader events will limit number of infections and lower the introduction of new variants and will also decrease the probability that a single large super-spreader event will initiate a more rapid local epidemic as has already occurred in Boston, South Korea and other locations during the pandemic (24, 25).

Our model has important limitations. While the model’s qualitative findings are robust, we cannot estimate the outbreak size necessary to ensure introduction of new variants into a population as many parameters required to do so are unknown. For instance, it is not yet clear whether the percentage of immunocompromised hosts varies across populations based on factors such as HIV prevalence and availability and use of immunosuppression for organ transplantation and cancer treatment. The number of secondary infections created by a person with new variants may also differ from that of other members of the population in ways that are difficult to project. On the one hand, these individuals may shed for longer and at a higher viral load (5, 7). Yet, they also may be more ill and therefore quarantined at home or in the hospital limiting contact exposures. Moreover, while all variants will be impacted in the same way by the introduction of NPIs such as masking and physical distancing, the utilization of these interventions varies considerably among regions and over time. Our model does not capture these nuances and in this sense is intended to be phenomenological only.

We demonstrate that new variants are frequently created and introduced into the population during large waves of SARS-CoV-2 infection. Yet, most variants ultimately burn out and those that ultimately predominate likely were associated with early super-spreader events. These variants are most likely to emerge when the previously dominant variant is decreasing.

## Methods

### SARS-CoV-2 within-host model

We used the within-host model describing the SARS-CoV-2 infection from our previous study (16). This model assumes that the contact of SARS-CoV-2 (V) with susceptible cells (S) produces infected cells at rate βVS which then generates new virus at a per-capita rate π. The model also incorporates the death of infected cells mediated by (1) the innate responses (δI^k^) and (2) the acquired immune responses 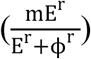 by SARS-CoV-2-specific effector cells (E). The magnitude of the innate immunity is dependent on the infected cell density and the exponent k. The nonlinearity of the acquired responses is captured by the Hill coefficient r that allows for rapid saturation of the killing. Finally, the parameter Φ defines level of SARS-CoV-2-specific effector cells at which the killing of infected cells becomes half maximal. In the model, the rise of SARS-CoV-2-specific effector cells rise is described in a two-stage manner. The first stage defines the proliferation of the first precursor cell compartment (M_1_) at rate ωIM_1_ and differentiation into a second precursor cell compartment (M_2_) at a per capita rate q. Finally, second precursor cells differentiate into effector cells at the same per capita rate q and are cleared at rate δ_E_.

The model is expressed as a system of ordinary differential equations:

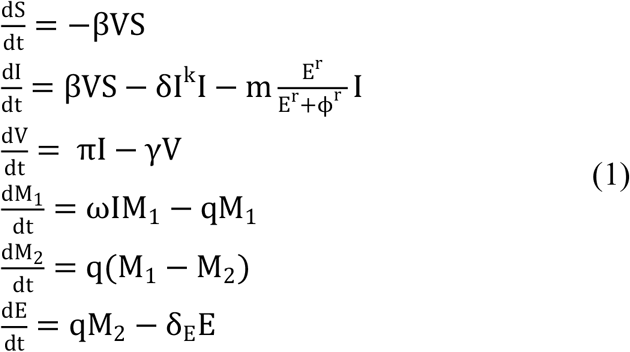

The initial conditions for the model were assumed as S(0) = 10^7^ cells/mL, I(0) = 1 cells/mL, 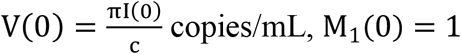 and E_0_ = 0. For simulations we sampled parameter values from a nonlinear mixed-effect model as described in (15), with the following fixed effects and standard deviation of the random effects (in parenthesis): Log_10_β: - 7.23 (0.2) virions^-1^ day^-1^; δ: 3.13 (0.02) day^-1^ cells^-k^; k: 0.08 (0.02); Log10(π): 2.59 (0.05) day^-1^; m: 3.21 (0.33) days^-1^cells^-1^; Log10(ω): -4.55 (0.01) days^-1^cells^-1^. We also assumed r = 10; δ_E_ = 1 day^-1^; q = 2.4 × 10^−5^ day^-1^ and c = 15 day^-1^.

### Dose-response model

We employed our previously developed dose-response model to estimate the probability of virus entering the airway given a transmitter viral load (i.e., contagiousness) and the probability of cellular infection given a transmitter viral load, (i.e., infectiousness) P_t_[V(t)] (response) based on viral loads V(t) (dose) (15). The relation between the response and the dose follows, 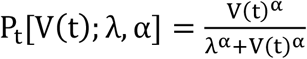, being λ the viral load that corresponds to 50% infectiousness and 50% contagiousness and α the Hill coefficient that controls the sharpness in the dose-response curve. We assumed that the viral load-dependent contagiousness (i.e., the probability that virus is passaged to the exposed person’s airway) is the same as infectiousness. We estimate the transmission risk as the product of the infectiousness and contagiousness (15).

### Transmission model and reproduction number

As in our previous model (26), we determined the total exposed contacts of a transmitter within a time step (Δ_t_) using a gamma distribution, i.e. 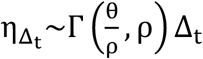, where θ and ρ represent the average daily contact rate and the dispersion parameter, respectively. The true number of exposure contacts (with viral airway exposure) was then obtained by multiplying the total exposed contacts and the contagiousness of the transmitter (i.e.,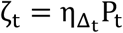). We modelled infectiousness as a Bernoulli event with mean P_t_, yielding the number of secondary infections within a time step as 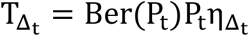. Finally, we summed up the number of secondary infections over 30 days since the time of exposure to obtain the individual effective reproduction number, i.e.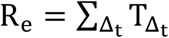. For each successful transmission, we further assumed that it takes τ days for the first infected cell to produce virus.

In simple steps, we followed the procedure below to estimate R_e_,

1. Simulate viral load V(t) of a simulated infected individual using the within-host model.
2. For a given combination of (λ, τ, α, θ, ρ)
  a. For each time step Δ_t_
    i. Compute P_t_[V(t); λ, α]
    ii. Draw 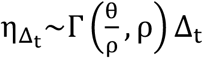
    iii. Calculate 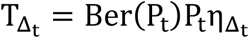
  b. Calculate 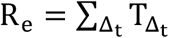
3. Repeat Steps 1 and 2 to estimate R_C_ for 1,000 infected individuals. The population level R_C_ can then be calculating by taking the mean of 1,000 individual R_C_ values.

### Parameter values for the transmission model

For simulations, we used the parameter set [α, λ, τ, θ, ρ] = [0.8, 10^7^, 0.5, 4, 40]) (15) as they most closely reproduces empirically observed individual R_e_ and serial interval histograms as well as mean R_e_ across individuals (R_0_ ∈ [1.4, 2.5]) and mean serial interval across individuals (SI ∈ [4.0, 4.5]) early during the pandemic (18, 19, 27-29).

### Simulating temporal dynamics from the transmission model

For a specific scenario with selected ρ and θ, we followed the procedure below to transform the transmission model into temporal transmission model:

1. Discretize the time-space of 150 days over time steps Δ_t_ of 1 day.
2. With n_t_ representing the number of transmitters at any time t, we start with presumed n_0_ transmitters at t=0 and zero transmitters at the remaining time points.
3. Starting at t = 0, we first determine the number of transmitters at that time step and then,
  a. For each of n_t_ transmitters:
    i. Simulate V(T) over [t, t + 30] at daily intervals (i.e., ΔT = 1) using the within-host model in eq. 1.
    ii. Compute P_T_[V(T); λ, α].
    iii. Draw 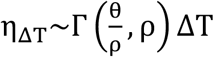.
    iv. Calculate 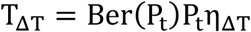.
    v. Determine times of successful transmission (t_S_) as those times ‘t’ where T_ΔT_ > 0 and the number of secondary transmissions at those time points as T_ΔT_.
    vi. Update 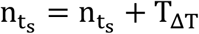.
4. Repeat Step 3 for t = Δ_t_, t = 2Δ_t_ and so on over the discretized time-space of 150 days.

### Simulating multi-class temporal dynamics from the transmission model

We assumed 7 classes of mutant strains, each with a different R_e_ of 1.0, 1.2, 1.4, 1.6, 1.8, 2.0 and 2.2. To simulate R_e_ of 1.0, 1.2, 1.4, 1.6, 1.8, 2.0 and 2.2, we employed θ of 2.3/day, 3.1/day, 3.5/day, 3.75/day, 4.0/day, 5.0/day and 5.5/day, respectively. For a specific scenario with selected ρ, θ and the probability µ of the transmitter transmitting mutant strain, we followed the procedure below to transform the transmission model into multi-class temporal transmission model:

1. Discretize the time-space of 150 days over time steps Δ_t_ of 1 day.
2. With n_tc_ representing the number of transmitters at any time t of class ‘c’, we start with presumed n_0c_ transmitters at t=0 of class c and zero transmitters at the remaining time points for all classes.
3. Starting at t = 0, for each of the seven classes,
  a. we determine the number of transmitters at that time step of class ‘c’ and then,
  b. For each of n_tc_ transmitters:
    i. Simulate V(T) over [t, t + 30] at daily intervals (i.e., ΔT = 1) using the within-host model in eq. 1.
    ii. Compute P_T_[V(T); λ, α].
    iii. Draw 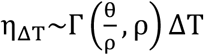.
    iv. Calculate 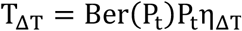.
    v. Determine times of successful transmission (t_S_) as those times ‘t’ where T_ΔT_ > 0 and the number of secondary transmissions at those time points as T_ΔT_.
    vi. Determine which strain was transmitted at times of successful transmission using µ_T_ = Ber(µ). If µ_T_ equals 1, then only a mutant strain is transmitted and the class of the mutant strain is randomly selected from 7 pre-specified classes.
    vii. Update n_tc_ = n_tc_ + T_ΔT_.
4. Repeat Step 3 for t = Δ_t_, t = 2Δ_t_ and so on over the discretized time-space of 150 days.

In the case of R_e_ = 0.8 (simulated with *θ* = 1.1/day), we assume 8 variants instead of 7 and follow steps 1-4 as mentioned above.

## Data Availability

All code is available upon request.

## References

1. N. G. Davies et al., Estimated transmissibility and impact of SARS-CoV-2 lineage B.1.1.7 in England. Science (2021).

2. H. Tegally et al., Emergence and rapid spread of a new severe acute respiratory syndrome-related coronavirus 2 (SARS-CoV-2) lineage with multiple spike mutations in South Africa. medRxiv, 2020.2012.2021.20248640 (2020).

3. N. R. Faria et al., Genomics and epidemiology of a novel SARS-CoV-2 lineage in Manaus, Brazil. medRxiv, 2021.2002.2026.21252554 (2021).

4. T. Bedford et al., Cryptic transmission of SARS-CoV-2 in Washington state. Science 370, 571–575 (2020).

5. B. Choi et al., Persistence and Evolution of SARS-CoV-2 in an Immunocompromised Host. N Engl J Med 383, 2291–2293 (2020).

6. A. L. Valesano et al., Temporal dynamics of SARS-CoV-2 mutation accumulation within and across infected hosts. bioRxiv (2021).

7. J. H. Baang et al., Prolonged Severe Acute Respiratory Syndrome Coronavirus 2 Replication in an Immunocompromised Patient. J Infect Dis 223, 23–27 (2021).

8. T. T. Truong et al., Persistent SARS-CoV-2 infection and increasing viral variants in children and young adults with impaired humoral immunity. medRxiv (2021).

9. N. G. Davies et al., Increased mortality in community-tested cases of SARS-CoV-2 lineage B.1.1.7. Nature (2021).

10. W. F. Garcia-Beltran et al., Multiple SARS-CoV-2 variants escape neutralization by vaccine-induced humoral immunity. Cell (2021).

11. S. A. Madhi et al., Efficacy of the ChAdOx1 nCoV-19 Covid-19 Vaccine against the B.1.351 Variant. N Engl J Med (2021).

12. S. Cele et al., Escape of SARS-CoV-2 501Y.V2 from neutralization by convalescent plasma. medRxiv, 2021.2001.2026.21250224 (2021).

13. E. C. Sabino et al., Resurgence of COVID-19 in Manaus, Brazil, despite high seroprevalence. Lancet (2021).

14. R. M. Coutinho et al., Model-based estimation of transmissibility and reinfection of SARS-CoV-2 P.1 variant. medRxiv, 2021.2003.2003.21252706 (2021).

15. A. Goyal, D. B. Reeves, E. F. Cardozo-Ojeda, J. T. Schiffer, B. T. Mayer, Viral load and contact heterogeneity predict SARS-CoV-2 transmission and super-spreading events. Elife 10, e63537 (2021).

16. A. Goyal, E. F. Cardozo-Ojeda, J. T. Schiffer, Potency and timing of antiviral therapy as determinants of duration of SARS-CoV-2 shedding and intensity of inflammatory response. Sci Adv 6, eabc7112 (2020).

17. A. Goyal, D. B. Reeves, E. F. Cardozo Ojeda, B. T. Mayer, J. T. Schiffer, Slight reduction in SARS-CoV-2 exposure viral load due to masking results in a significant reduction in transmission with widespread implementation. medRxiv, 2020.2009.2013.20193508 (2020).

18. A. Endo, S. Abbott, A. J. Kucharski, S. Funk, C. f. t. M. M. o. I. D. C.-W. Group, Estimating the overdispersion in COVID-19 transmission using outbreak sizes outside China. Wellcome Open Res 5, 67 (2020).

19. Q. Bi et al., Epidemiology and transmission of COVID-19 in 391 cases and 1286 of their close contacts in Shenzhen, China: a retrospective cohort study. Lancet Infect Dis 20, 911–919 (2020).

20. H. L., D. P., C. I., e. al., High SARS-CoV-2 Attack Rate Following Exposure at a Choir Practice — Skagit County, Washington, March 2020.. MMWR Morb Mortal Wkly Rep 69:606–610. (2020).

21. J. Brugger, C. L. Althaus, Transmission of and susceptibility to seasonal influenza in Switzerland from 2003 to 2015. Epidemics 30, 100373 (2020).

22. S. Tang et al., Aerosol transmission of SARS-CoV-2? Evidence, prevention and control. Environ Int 144, 106039 (2020).

23. P. Bittihn, R. Golestanian, Stochastic effects on the dynamics of an epidemic due to population subdivision. Chaos 30, 101102 (2020).

24. J. E. Lemieux et al., Phylogenetic analysis of SARS-CoV-2 in Boston highlights the impact of superspreading events. Science 371 (2021).

25. https://www.cdc.go.kr/board/board.es?mid=a30402000000&bid=0030&act=view&list_no=366485&tag=&nPage=1(

26. A. Goyal, D. B. Reeves, E. F. Cardozo-Ojeda, J. T. Schiffer, B. T. Mayer, Wrong person, place and time: viral load and contact network structure predict SARS-CoV-2 transmission and super-spreading events. medRxiv, 2020.2008.2007.20169920 (2020).

27. Y. Zhang, Y. Li, L. Wang, M. Li, X. Zhou, Evaluating Transmission Heterogeneity and Super-Spreading Event of COVID-19 in a Metropolis of China. Int J Environ Res Public Health 17 (2020).

28. Z. Du et al., Serial Interval of COVID-19 among Publicly Reported Confirmed Cases. Emerg Infect Dis 26 (2020).

29. D. Adam et al., Clustering and superspreading potential of severe acute respiratory syndrome coronavirus 2 (SARS-CoV-2) infections in Hong Kong. Europe PMC 10.21203/rs.3.rs-29548/v1 (2020).

